# Study of the protective *OAS1* rs10774671-G allele against severe COVID-19 in Moroccans suggests a North African origin for Neanderthals

**DOI:** 10.1101/2023.08.19.23294314

**Authors:** Fatima Zohra El Youssfi, Abbas Ermilo Haroun, Chaimae Nebhani, Jihane Belayachi, Omar Askander, Elmostafa El Fahime, Hakima Fares, Khalid Ennibi, Redouane Abouqal, Rachid Razine, Ahmed Bouhouche

## Abstract

**Background:** The clinical presentation of COVID-19 has shown high variability between individuals, which is partly due to genetic factors. The OAS1/2/3 cluster was found to be strongly associated with COVID-19 severity. We aimed to examine this locus for the occurrence of the critical variant, rs10774671, and its respective haplotype blocks within the Moroccan population.

**Methods:** The frequency of SNPs at the cluster of OAS immunity genes was assessed from an in-house database in 157 unrelated individuals of Moroccan origin. The *OAS1* exon 6 was sequenced by Sanger’s method in 71 asymptomatic/mild and 74 moderate/severe individuals positive for SARS-CoV-2. Genotypic, allelic, and haplotype frequencies of three SNPs were compared between the two groups. Finally, males in our COVID-19 series were genotyped for the Berber-specific marker E-M81.

**Results:** The prevalence of the *OAS1* rs10774671-G allele in present-day Moroccans was 40.4%, close to that of Europeans. However, it was found equally on both the Neanderthal GGG haplotype and the African GAC haplotype with a frequency of 20% each. These two haplotypes, and hence the rs10774671-G allele, were significantly associated with the protection against severe COVID-19 (*p* = 0.034, *p* = 0.041, and *p* = 0.008 respectively). Surprisingly, among Berber men, the African haplotype was absent while the prevalence of the Neanderthal haplotype was close to that of Europeans.

**Conclusion:** The protective rs10774671-G allele of *OAS1* was found only in the Neanderthal haplotype in Berbers, the indigenous people of North Africa, suggesting that this region may have served as the stepping-stone for the passage of the hominids to the other continents.

## 1. Introduction

COVID-19, caused by the coronavirus SARS-CoV-2, appeared in November 2019 in Wuhan, Hubei province (China), before spreading around the world and causing a pandemic [1, 2]. The consequences of SARS-CoV-2 infection vary greatly from person to person and from population to population. While most infected individuals are asymptomatic or minimally symptomatic, some develop severe or even critical illness. Older people and those with underlying medical condition, such as cardiovascular disease, diabetes, chronic respiratory disease, or cancer, are at greater risk of developing a severe form [3]. However, these risk factors alone do not explain the variability in severity observed. These differences could partly result from individual genetic susceptibility. The scientific community has been mobilized to respond to the health crisis to identify the genetic determinants of COVID-19 that protect people from or predispose them to severe manifestations of the disease. A genetic study conducted on patients of European ancestry identified a risk locus for severe COVID-19 in the 2’-5’-oligoadenylate synthetase 1/2/3 (OAS1/2/3) gene cluster [4]. Subsequently, it was reported that elevated levels of OAS1 in the plasma were associated with reduced COVID-19 susceptibility and severity [5].

This genetic region of chromosome 12q24.13 carries a haplotype of approximately 75 kb of Neanderthal origin [6], where the discovery of the causal variant was possible through studies conducted on a multiancestry cohort including patients from both African and European ancestry. It consists of rs10774671 with G>A transition occurring at a splice acceptor site, where the protective ancestral G allele produces a longer OAS1 transcript and a more active protein [7, 8]. Moreover, *OAS1* SNPs have been reported to be associated with susceptibility to West Nile virus infections [9] and to be involved in altered cellular function leading to several autoimmune and infectious diseases, suggesting that the *OAS* genes play a critical role in the innate immune response to viruses [10–14]. For instance, the OAS1 enzyme catalyzes the synthesis of short polyadenylates, which activate ribonuclease L contributing to the degradation of viral RNA and inhibition of viral replication [15, 16]. It produces five transcripts including p42, p44, p46, p48, and p52, and its expression is stimulated by the interferon system [17, 18].

In this study, we assessed the prevalence of several variants at the *OAS* gene cluster that comprise the Neanderthal haplotype by analyzing exome data of 157 unrelated Moroccan individuals and observed that the *OAS1* rs10774671-G allele was found on both Neanderthal and African haplotypes. We then investigated the rs10774671 variant in 146 Moroccan volunteers positive for SARS-CoV-2 infection and showed that it was significantly associated with COVID-19 severity. Interestingly, in Berbers of both paternal and maternal lineages, the African haplotype containing the G allele was absent while the frequency of Neanderthal-derived haplotypes is close to that of Europeans.

## Material and methods

### Frequency of *OAS* gene variants in the Moroccan population

To assess the frequency in the Moroccan population of the Neanderthal haplotype at the cluster of *OAS* immunity genes in the chromosomal region 12q24.13, we analyzed exome data from 157 unrelated patients suspected of having a rare hereditary disease. These patients were recruited from the Department of Neurology of Specialties Hospital and Cheikh Zaid Hospital of Rabat. Part of the data was taken from the in-house exome database and the other part comes from whole exome sequencing (WES) carried out as part of a collaborative project with 3Billion® (Seoul, Korea). Data were collected for variants rs1131454, rs10774671, rs1131476, rs1051042 and rs2660 in *OAS1*, rs1859330, rs1859329 and rs2285932 in *OAS3*, and the variant rs1293767 in *OAS2*, variants that comprise the Neanderthal haplotype at the *OAS* locus as described elsewhere [19].

### COVID-19 Patients recruitment

To highlight whether the variants studied are associated with the severity of COVID-19 in the Moroccan population, it was essential to minimize the effects of underlying medical conditions such as comorbidities by reducing the upper age limit of patients. Thus, we have adopted the selection criteria including males or females of Moroccan origin, aged between 18 and 65 years old, with positive RT-PCR test for SARS-COV-2, who have not received any vaccines. Pregnant patients were excluded from the study. 146 COVID-19 patients were enrolled for 1 year between 28 July 2020 to 15 July 2021, a period of the pandemic dominated by the alpha and beta variants. All patients filled out a form concerning socio-demographic data for the establishment of ethnicity and medical history, notifying comorbidities, if applicable. Patients were recruited by three different hospital centers involved in a national COVID-19 research consortium, including the Medical Emergency Department (CHU Ibn Sina, Rabat), the Virology Department (Military Hospital, Rabat), and the Intensive Care Unit (Cheikh Zaid international university hospital, Rabat). Outcomes were defined as asymptomatic (without symptoms), mild (non-hospitalized), moderate (hospitalized but not requiring invasive ventilation), and severe (hospitalized requiring invasive ventilation or death). 146 patients were recruited and grouped into the following severity categories, asymptomatic (N = 21), mild COVID-19 (N = 50), moderate COVID-19 (N = 48), and severe COVID-19 (N = 27). Asymptomatic and mild patients were grouped (N=71) and compared to moderate/severe patients (N=75). Five ml of venous blood was collected in an EDTA tube from each participant and stored at −20°C until DNA extraction. The written informed consent was signed by all patients before their enrollment. This study was approved by the ethics committee of Cheikh Zaid Hospital of Rabat (ID: CEFCZ/AB/PR_RFG).

### Genetic analysis

Genomic DNA (gDNA) of 146 SARS-CoV-2 positive volunteers was extracted using the MagPurix® Blood DNA Extraction Kit with a Zinexts MagPurix EVO 24 CE IVD system (Zinexts Life Science Corp., New Taipei City, Taiwan). Quality and concentrations of DNA samples were analyzed by Nanodrop One^c^ Spectrophotometer (Thermo Fisher Scientific). PCR amplification, covering the *OAS1* exon 6 and its intron boundaries, was performed using two pairs of specific primers (forward: 5’-GTGGCCAGGCTTCTATACCC-3’; and reverse: 5’-TGGAGTGTGCTGGGTCTATG-3’) designed with Primer3 Input (V 0.4.0). PCR amplification was performed using 100 ng of template gDNA, 1 µL of each primer (10 µM), 5 µL of 5X MyTaq reaction buffer, and 0.2 µl of MyTaq DNA polymerase (Bioline) in a total reaction volume of 25 µL. Thermal conditions were as follows: 95°C for 3 min, 35 cycles of 95°C for 5 sec, 60°C for 15 sec, and 72°C for 10 sec, with a final extension of 72°C for 10 min. The PCR products were visualized by 1.5% agarose gel electrophoresis. Five microliters of PCR amplicons were purified using ExoSap IT-express reagent and 2 microliters of the product were used for cycle sequencing with BigDye™ Terminator v3.1 Ready Reaction Cycle Sequencing Kit. Electrophoresis was performed on SeqStudio Genetic Analyzer, and Sequence data were analyzed by SeqScape 4 software (Thermo Fisher Scientific).

The genetic marker, E-M81, reported as being the specific male lineage of autochthonous Berbers of North Africa, was analyzed in all 80 males in the COVID-19 positive cohort of patients. The marker was amplified by PCR and Sanger sequenced as described previously [20].

### Statistical analysis

Statistical analysis was performed using *jamovi* software (Version 1.6). The evaluation of demographic variables and disease characteristics was done using descriptive statistics. A chi-square (χ2) test was used before the association tests to assess deviation from Hardy-Weinberg equilibrium (HWE) in the control group. The genotypic and allelic frequencies were compared between patients and controls to check the associations of the studied SNPs with COVID-19 severity. The χ2 test was used to identify the differences in genotypic distribution between study groups under an additive genetic model. For allelic and haplotype models, Odds ratios (ORs) at 95% confidence intervals (CI) were estimated using logistic regression analysis, and *p*-values less than 0.05 was considered statistically significant.

## Results

### Frequency of Neanderthal alleles at *OAS* gene cluster in Moroccans

The genotype of 9 SNPs at the cluster of *OAS* immunity genes in the chromosomal region 12q24.13 (Fig 1) was extracted from an exome sequencing database of 157 unrelated individuals, all of Moroccan origin. The results presented in Table 1 showed that, overall, the frequencies of these SNPs in Moroccans are close to the frequency of the minor allele frequency (MAF) of the 1000 genome project. Focusing on *OAS1* variants, the reference G allele of the *OAS1* splice variant rs10774671 represented 40.4% in analyzed Moroccan individuals, close to that of Europeans with 35.2%, but much lower than that of Africans with 64.0% and much higher than that of Asians with only 27.2%. The frequencies of the missense variants rs1131476 and rs1051042, and the 3’UTR rs2660 variant were 21.1% identical, indicating that they are in complete linkage disequilibrium, which was also the case in the other populations of European, Asian and African ancestry (Table 1).

**Fig 1:**
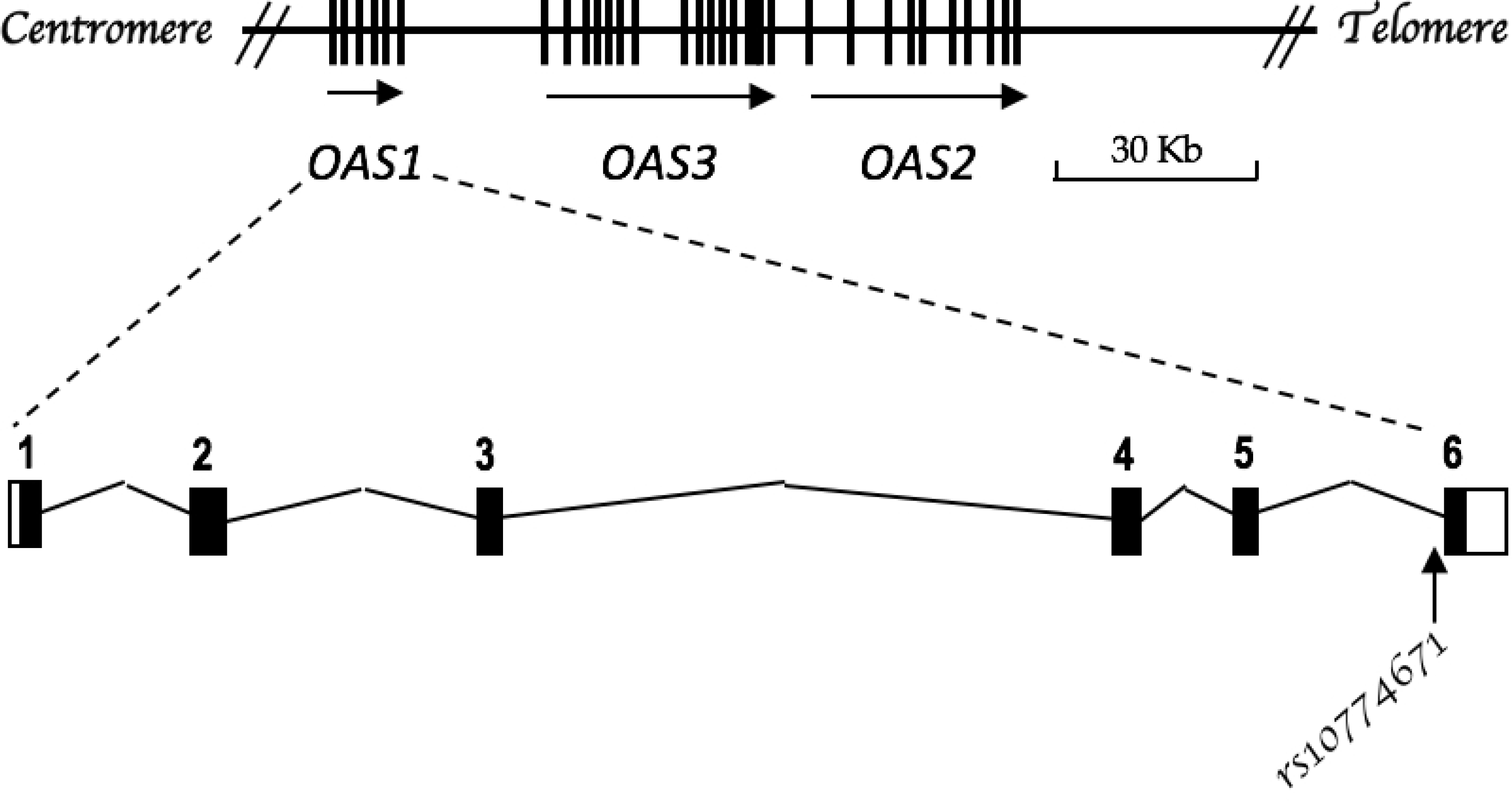
OAS gene cluster at chromosome 12q24.13 and structural organization of OAS1 giving the location of the splice variant rs10774671.

**Table 1:** SNP Frequencies of the OAS locus in 157 individuals of Moroccan origin in comparison with those of the 1000 genomes project.

|  | **OAS1** |  |  |  |  | **OAS3** |  |  | **OAS2** |
| --- | --- | --- | --- | --- | --- | --- | --- | --- | --- |
|  | rs1131454<br>c.484G>A<br>p.Gly162Ser | rs10774671<br>c.1039-1G>A<br>Splice site | rs1131476<br>c.1054G>A<br>p.Ala352Pro | rs1051042<br>c.1082G>C<br>p.Arg361Met | rs2660<br>c.*84G>A<br>3'UTR | rs1859330<br>c.53G>A<br>p.Arg18Lys | rs1859329<br>c.117C>T<br>p.Ala39= | rs2285932<br>c.1314T>C<br>p.Ile438= | rs1293767<br>c.448+41C>G<br>Intronic |
| Minor allele frequency | 0.470 | 0.390 | 0.210 | 0.210 | 0.210 | 0.340 | 0.210 | 0.160 | 0.150 |
| African Ancestry | 0.851 | 0.640 | 0.027 | 0.027 | 0.027 | 0.446 | 0.011 | 0.010 | 0.011 |
| Asian Ancestry | 0.414 | 0.272 | 0.271 | 0.271 | 0.271 | 0.268 | 0.268 | 0.156 | 0.130 |
| European Ancestry | 0.425 | 0.352 | 0.345 | 0.345 | 0.345 | 0.370 | 0.365 | 0.328 | 0.325 |
| **Moroccan population** | **0.554** | **0.404** | **0.210** | **0.210** | **0.210** | **0.283** | **0.213** | **0.197** | **0.194** |

We then explored the haplotypes formed by the three SNPs rs10774671, rs1131476, and rs1051042, which allowed for identification of the predominantly protective Neanderthal-derived haplotype in this genetic region. The haplotypes consisting of these three SNPs were GGG, GAC, and AAC, and the graphical representation of their distribution in the present-day Moroccans in comparison with different ancestries are shown in Fig 2. The *OAS1* rs10774671-G allele in Moroccans was included in both Neanderthal GGG and African GAC haplotypes with frequencies of 21% and 19.4% respectively. In comparison to other ancestries, the GGG haplotype accounts for 34.5% of Europeans, 27.1% of Asians, and only 2.7% of Africans. The GAC haplotype is however predominant in Africans with a frequency of 61.3% and is almost absent in Europeans and Asians with 0.7% and 0.001% respectively. Finally, the risk haplotype AAC was the prevalent haplotype in the Asian population with 72.4% (Table 1).

**Fig 2.**
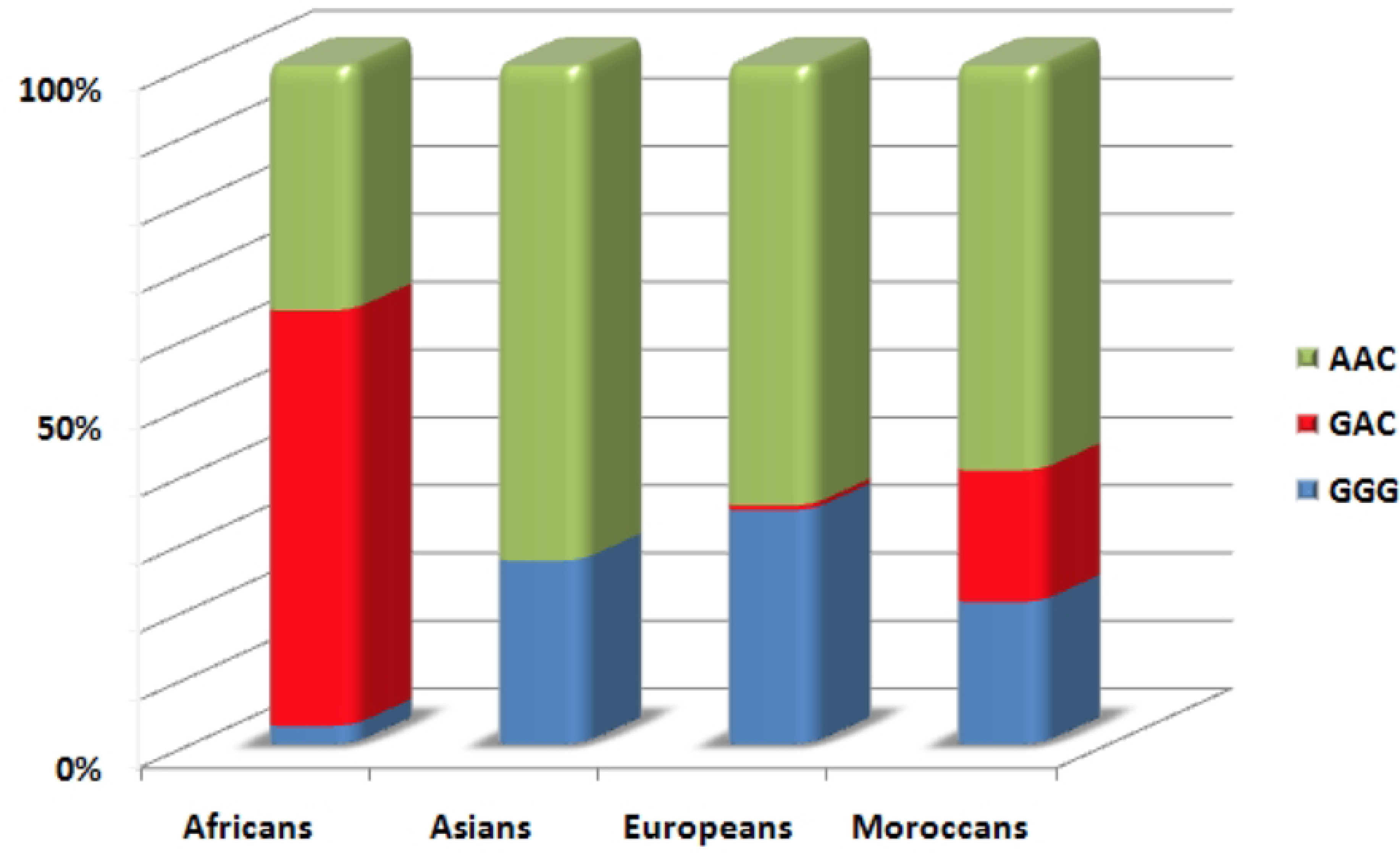
Prevalence of the three possible haplotypes with the three OAS1 SNPs studied in 157 individuals of Moroccan origin in comparison with the data of 1000 Genome Project of Africans, Asians and Europeans.

### Association study in Moroccan COVID-19 patients

The demographic and clinical data of the two studied groups are shown in Table 2. The mean age and standard deviation of patients asymptomatic/mild were 44.1 ± 13.5 years and significantly lower than that of moderate/severe with 51.4 ± 10.5 years (*p* < 0.001). There was no significant difference regarding the sex ratio between the two groups (OR = 1.04, *p* < 0.897), with a female-to-male ratio of approximately 1:1. As expected, the patients in the asymptomatic/mild group displayed a significantly lower frequency of comorbidity compared to the moderate/severe group (OR = 2.85, p < 0.004), with 22.5% and 56%, respectively. Likewise, the mortality rate was 0% among the asymptomatic/mild patients and 8% among moderate/severe patients, which was significantly different (OR = 0.07, *p* < 0.015).

**Table 2:** Demographic criteria of COVID-19 studied groups.

|  | Disease severity |  |  |  | OR (CI 95%) | *p* value |
| --- | --- | --- | --- | --- | --- | --- |
|  | Asymptomatic/Mild |  | Moderate/Severe |  |  |  |
|  | (n = 71) |  | (n = 75) |  |  |  |
|  | No | % | No | % |  |  |
| Age (y) Mean ± SD | 44.1 + 13.5 |  | 51.4 + 10.5 |  |  | < .001 |
| Sex |  |  |  |  |  |  |
| Male | 39 | 54.9 | 41 | 54.7 | 1.04 (0.544-2.01) | 0.897 |
| Female | 32 | 45.1 | 34 | 45.3 |  |  |
| Comorbidity | 16 | 22.5 | 42 | 56.0 | 2.85 (1.39-5.85) | 0.004 |
| Mortality | 00 | 0.0 | 06 | 8.0 | 0.07 (0.004-1.35) | 0.015 |

The distribution of genotypes, alleles, and haplotypes of the three *OAS1* SNPs according to severity were presented in Table 3. Among the 292 alleles analyzed, the ancestral G allele represented 40% for the rs10774671variant and 21.23% for both the rs1131476 and rs1051042 variants. The comparison between the two groups studied, asymptomatic/mild versus moderate/severe, shows a significant difference under both the additive (*p* = 0.014) and allelic models (OR 0.528 [95% CI 0.329-0.848], *p* = 0.008). However, no statistically significant differences in the distribution of the rs1131476 polymorphism were observed between the two study groups under both the additive (*p* = 0.248), and allelic models (OR 0.617 [95% CI 0.350-1.090], *p* = 0.096), suggesting that this missense mutation was not associated with COVID-19 severity in Moroccans.

**Table 3.**
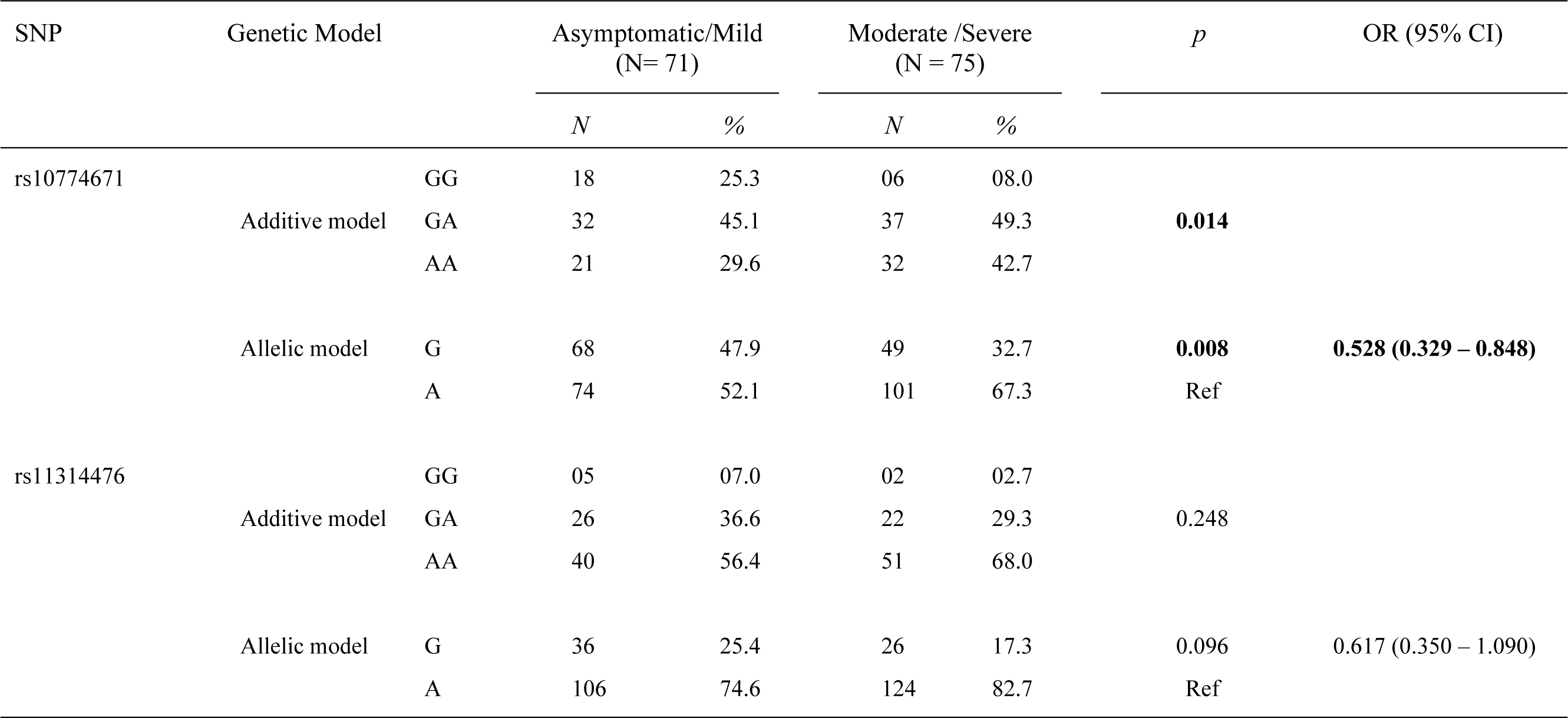
Comparison of genotypic and allelic distribution of the OAS1 rs10774671 and rs11314476 SNPs between COVID-19 asymptomatic/mild and Moderate/Severe groups.

The comparison of the occurrence of these polymorphisms in their respective haplotype block between the two groups studied is shown in Table 4. The three selected SNPs formed only three haplotypes GGG, GAC and AAC. The two haplotypes GGG, and GAC were common in Asymptomatic/Mild versus Moderate/Severe patients and associated compared to the risk AAC haplotype with OR 0.529 [95% CI 0.285 – 0.973], *p* = 0.034 and OR 0.527 [95% CI 0.028 – 0.509], *p* = 0.041 respectively. These two haplotypes thus confer protection against severe COVID compared to the risk haplotype AAC in patients of Moroccan origin.

**Tables 4.** The comparison of the haplotype distribution consisting of rs10774671, rs1131462 and rs1051042 between the asymptomatic/mild group versus moderate/severe group.

|  | Asymptomatic/Mild<br>(N= 71) |  | Moderate /Severe<br>(N = 75) |  | *p* | OR (95% CI) |
| --- | --- | --- | --- | --- | --- | --- |
|  | *N* | % | *N* | % |  |  |
| Haplotype |  |  |  |  |  |  |
| GGG | 36 | 25.4 | 26 | 17.3 | **0.034** | **0.529** (0.285 – 0.973) |
| GAC | 32 | 22.5 | 23 | 15.3 | **0.041** | **0.527** (0.028 – 0.509) |
| AAC | 74 | 52.1 | 101 | 67.4 | Ref |  |

### Distribution of *OAS1* haplotypes in Berbers

Among the 80 males in our cohort of SARS-CoV-2-positive individuals, 41 were positive and 39 were negative for the E-M81 marker. The distribution of the three haplotypes studied GGG, GAC and AAC in these two groups, as well as that of 29 males with Parkinson’s disease carrying the *LRRK2* G2019S mutation whose Berber origin has been established for both paternal and maternal lineages [20] were presented in Fig 3. The results showed that the prevalence of the African GAC haplotype in males with the E-M81 marker represented only 8.5% and was lower than that of males negative for this marker with 25.7%. Furthermore, this GAC haplotype was completely absent in males whose Berber origin concerned both the maternal and paternal lineages. The prevalence of both GGG and AAC haplotypes in these pure Berber males is close to that of European ancestry.

**Fig 3.**
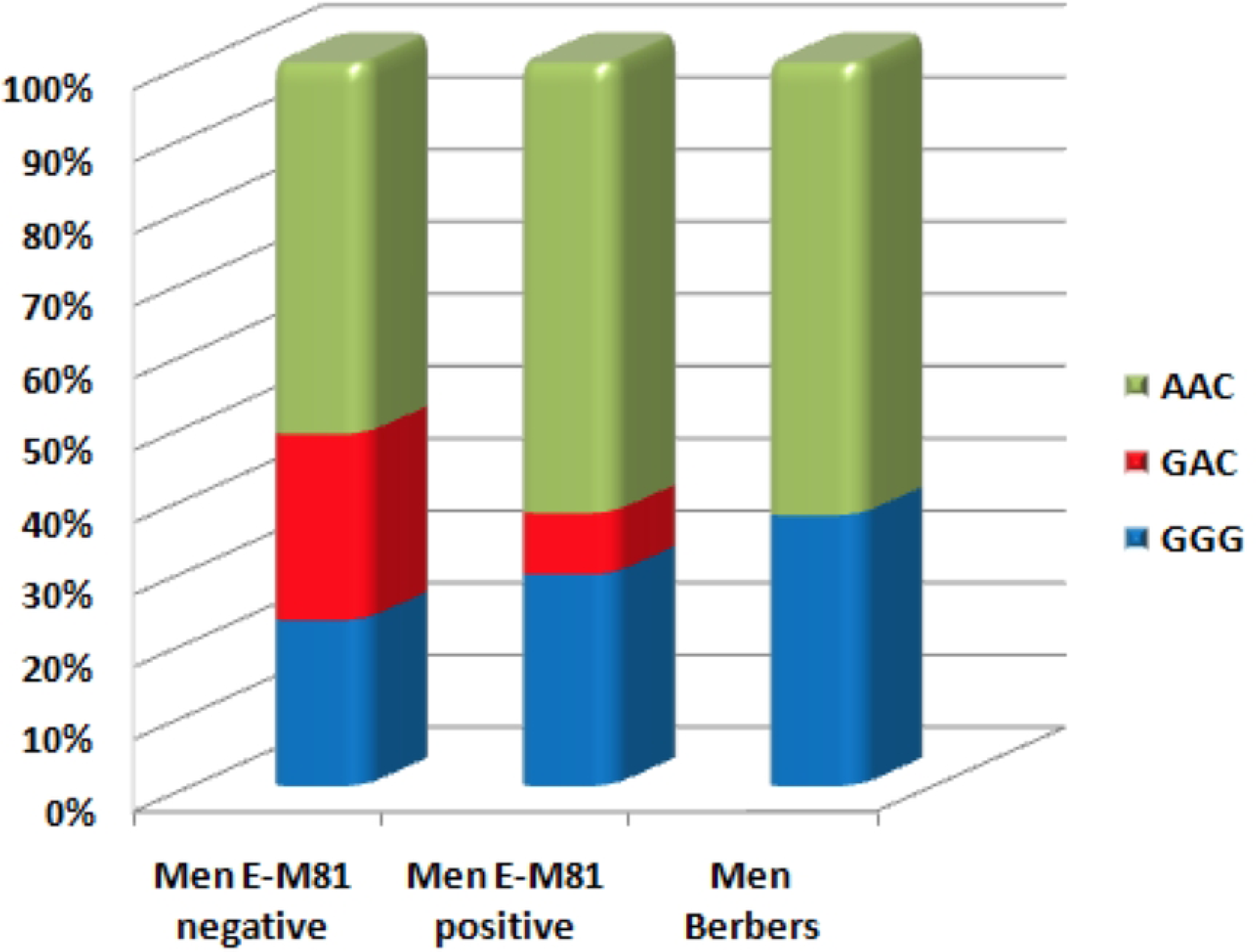
Frequencies of the three haplotypes consisting of the three OAS1 SNPs studied in 39 males negative and 41 males positive for E-M81 marker, all positive for SARS-CoV-2, and 29 males *LRRK2* G2019S carriers whose Berber origin has been established for both paternal and maternal lineages.

## Discussion

The clinical presentation of COVID-19 following SARS-CoV-2 infection has shown high interindividual variability, ranging from asymptomatic to death, which is partly due to genetic factors. The chr12q24.13 region containing the *OAS* gene cluster was recently reported to be strongly associated with COVID-19 severity and mortality [4], and the rs10774671 splice variant was confirmed to be the functionally critical variant of this chromosomal region that influences the outcomes of SARS-CoV-2 infection [7, 8]. Indeed, the *OAS1* rs10774671-G allele has been shown to increase levels of the p46 isoform and thus the circulating levels of OAS1 protein that were strongly associated with reduced risks of severe COVID-19 [5]. This protective G allele represents about 35% of individuals of European ancestry and was found on a haplotype inherited from the Neanderthal, and it represents 64% of the African population independently of this Neanderthal-derived haplotype [6].

To our knowledge, no data currently exist on the prevalence of *OAS* gene cluster variants in the North African population, which by its geographical position in the African continent could give relevant information on how this G allele at rs10774671 was inherited from the ancestral population common to both modern humans and Neanderthals. Our study thus presents a first insight into this chromosomal region in the Moroccan population, which is culturally, historically, and anthropologically similar to the other neighboring countries of the North African subcontinent. We showed that the frequencies of *OAS* cluster SNPs in Moroccans were close to the MAF of the 1000 genome project, where the *OAS1* rs10774671-G allele represented 40.4%. However, analyzing the haplotypes consisting of three *OAS1* variants that captured the Neanderthal haplotype showed that the rs10774671-G allele was found equally on both the Neanderthal GGG haplotype and the African GAC haplotype. This distribution seems different from the European population where the G allele was mostly found at 35% in the Neanderthal haplotype, and from the African population where it was mostly found at 64% in a specific African haplotype, as reported before [6–8].

The study of these 3 selected SNPs in a series of 146 SARS-CoV-2 positive individuals of Moroccan origin less than 65 years of age showed that only the splice variant rs10774671 presents a significant association with the severity of COVID-19. Both the AA genotype and the A allele were predictive of severe COVID-19 disease, supporting literature findings that this is the only variant of this locus influencing COVID-19 outcomes [7, 8]. However, even though the prevalence of the protective allele *OAS1* rs10774671-G in the Moroccan population was close to that of Europeans, the severity and mortality caused by SARS-CoV-2 infection are much higher in individuals of European ancestry, even leading to a reduction in life expectancy [21]. This suggests that other genetic factors as well as general health factors and conditions, all interacting in complex ways, could be differentially involved between the two populations and remain to be discovered.

Furthermore, both the Neanderthal GGG and African GAC haplotypes, containing the rs10774671-G allele were shown to be protective against severe COVID-19. Overall, in the 146 individuals positive for SARS-CoV-2, the frequency of the G allele and of the two GGG and GAC haplotypes were consistent with those obtained in the control cohort of our WES database, thus confirming the equal prevalence of around 20% for each of the two Neanderthal and African haplotypes in the present-day Moroccans. The presence of the African haplotype could certainly be due to a recent admixture with sub-Saharan African populations, while the chronology of the presence of the Neanderthal haplotype in this population would be difficult to determine precisely.

The Moroccan population is known to be a heterogeneous population having received influences from the south, north, and east, and whose ancient origin was essentially indigenous Berber. To provide elements of answers to the hypothesis of the origin of the Neanderthal Haplotype in North Africa, the distribution of the three haplotypes was compared in our series of SARS-CoV-2 positive individuals between males with and without the marker E-M81, known to be the specific male lineage of autochthonous Berbers of North Africa. The frequencies of the African GAC haplotype decreased to 8.5% in favor of the Neanderthal GGG haplotype which increased to 29.3% in males with the E-M81 marker compared to males without this marker at a frequency of 23% and 25.7% respectively. GAC haplotypes in males with the E-M81 marker are likely derived from maternal lineage. Surprisingly, the African GAC haplotype was completely absent in Berber males from both paternal and maternal lineages, and whose frequencies of the two remaining haplotypes GGG and AAC were similar to those of European ancestry.

These results provide evidence for the presence of the Neanderthal protective haplotype among Berbers who are the ancient indigenous inhabitants of North Africa over the last 10,000 years. However, the question that remains to be asked is whether this haplotype was imported to North Africa from the “back to Africa” migration or exported to Europe and Asia by the earlier “out of Africa” one. Several recent pieces of scientific evidence lean towards the second scenario. First, it has been reported that present-day North Africans share a majority of their ancestry with Europeans and West Asians but not with sub-Saharan Africans [22–24]. Along the same lines and in a slightly earlier period, the genetic study of human fossils from the Taforalt region dating back 15,000 years excluded gene flow from Southern Europe into northern Africa, as well as a genetic affinity with early Holocene Near Easterners, suggesting a late Pleistocene connection between North Africa and the Near East [25]. However, due to the absence in this region from ancient genomic data at a similar time, the authors could not define the epicenter or the direction of this connection.

The study of the G2019S mutation of the *LRRK2* gene, a founder-effect mutation found in the haplotype 1 identified worldwide but with varying prevalence, was previously determined to originate from a Near-Eastern founder at least 4000 years ago and reintroduced by recurrent gene flow to European and North-African populations [26]. Using uniparental markers, we showed that the mutation arose in a Berber founder and its epicenter was indeed in North Africa, where the prevalence was the highest with 40%, and the gene flow by migration took place in the opposite direction [20].

In the same way, the site of Jebel Irhoud located in Morocco has provided valuable information missing in the puzzle of the history of hominids and in particular their first exit from Africa to other continents. Irhoud hominids were originally considered as North African *H. sapiens* interbred with Neanderthals, an African form of Neanderthals, or a North African archaic population [27]. Recently, Hublin et al. [28] thermoluminescently dated the Irhoud fossils to 315,000 years ago, making Jebel Irhoud the oldest African hominid site with features of anatomically modern humans and more primitive cranial morphology. Phylogenetic modeling of cranial morphology showed phenotypic affinities of Irhoud fossils with both Neanderthals and early *H. sapiens* suggesting that they have been introgressed into Neanderthals at the end of the Middle Pleistocene contributing to the evolution of the Neanderthals [29]. Finally, we performed a whole genome sequencing of only three present-day Moroccans and identified over 200,000 SNV absent from 1000G and gnomAD databases, suggesting that the Moroccan population would have more genetic variability and therefore would be older than the European population. Principal components analysis showed that Moroccan genomes form a distinct population and placed them between European and African 1000G populations [30].

Sánchez-Quinto et al. [31] analyzed 780,000 SNPs in individuals from different locations in North Africa including Morocco and showed that North African populations have a significant excess of derived alleles shared with Neanderthals, whereas Sub-Saharan populations are not affected by this admixture event. The authors also showed that the Neanderthal’s genetic signal was higher in populations with a local, pre-Neolithic North African ancestry, confirming that this detected ancient admixture is not due to recent Near Eastern or European migrations. Furthermore, the first discovered Neanderthal fossil was at the Jbel Irhoud site [32], where Jean-Jacques Hublin et al. [28] dated the fossils found at 300,000 years old, that are accompanied by a Mousterian-type lithic industry, a lithic culture that has long been associated only with Neanderthals in Eurasia. Lithic data (Mousterians) have been reported long before this, and the oldest lithic remains discovered in Morocco belong to the Acheulean and are dated to around 1.3 million years [33, 34]. These data therefore constitute genomic, fossil and lithic evidence of the existence of Neanderthals in Morocco and their local contributions to anatomically modern humans, reinforcing the idea of a local introgression of the *OAS1* rs10774671-G allele.

Admittedly, Africa is the cradle of humanity, but the evolution of the hominid ancestors of *Homo Sapiens* did not occur in a linear way within this continent, but rather, through a focus dictated by severe geographical and climatic conditions. Therefore, the evolution of hominids in North Africa, isolated to the south by a largely arid and desert region, would have been different from that of sub-Saharan Africa, and would probably be at the origin of the initial dispersal out of Africa. Their conquest of the other continents would have been possible via the Strait of Gibraltar during the Ice Age, and later along the Mediterranean via the Fertile Crescent. The answers to this scenario are most likely to be found in the whole genome sequencing of a large series of present-day individuals and also in hominid fossils from North Africa.

## Conclusion

The rs10774671-G allele of *OAS1* confers protection against severe COVID-19 in the Moroccan population. Surprisingly, this protective allele was found in the Neanderthal haplotype in Berbers, the indigenous people of North Africa suggesting that this subcontinent could be the stepping-stone for the passage of the hominids to other continents.

## Supporting information

**S1 Table. Demographic, COVID-19 phenotype, genotype and haplotype of 3 OAS1 SNP of 146 individuals positive for SARS-COV-2.** (xlsx)

**S1 Table. Genotype of Y-M81 marker in males positive for SARS-COV-2.** (xlsx)

## Acknowledgments

We thank all the participants in this study, and 3billion® (Seoul, Korea) for providing part of the whole exome sequencing free of charge.

## Author Contributions

**Conceptualization:** Ahmed Bouhouche.

**Formal analysis:** Rachid Razine.

**Data Curation:** Omar Askander, Elmostafa El Fahime, Ahmed Bouhouche.

**Funding acquisition:** Ahmed Bouhouche.

**Investigation:** Chaimae Nebhani, Fatima-Zahra El Youssfi, Ahmed Bouhouche, Redouane Abouqal, Khalid Ennibi, Hakima Fares, Abbas Ermilo Haroun, Jihane Belayachi.

**Methodology:** Ahmed Bouhouche.

**Project administration:** Ahmed Bouhouche.

**Supervision:** Ahmed Bouhouche.

**Validation:** Ahmed Bouhouche.

**Writing – review & editing:** Ahmed Bouhouche.

## Data availability statement

All relevant data are within the manuscript and its Supporting Information files.

